# Seq-ing the SINEs of Central Nervous System Tumors in Cerebrospinal Fluid DNA

**DOI:** 10.1101/2022.06.28.22276835

**Authors:** Christopher Douville, Samuel Curtis, Mahmoud Summers, Tej D. Azad, Jordina Rincon-Torroella, Bracha Avigdor, Jonathan Dudley, Joshua Materi, Divyaansh Raj, Sumil Nair, Debarati Bhanja, Kyle Touhy, Lisa Dobbyn, Maria Popoli, Janine Ptak, Nadine Nehme, Natalie Silliman, Cherie Blair, Kathy Judge, Gary L. Gallia, Mari Groves, Christopher M. Jackson, Eric M. Jackson, John Laterra, Michael Lim, Debraj Mukherjee, Jon Weingart, Jarushka Naidoo, Carl Koschmann, Natalya Smith, Karisa C. Schreck, Carlos A. Pardo, Michael Glantz, Matthias Holdhoff, Kenneth W. Kinzler, Nickolas Papadopoulos, Bert Vogelstein, Chetan Bettegowda

## Abstract

Lesions within the brain cavity pose critical challenges for diagnostics, particularly distinction between cancerous and non-cancerous lesions. We here introduce an analytic technique called Real-CSF to detect cancers of the central nervous system from evaluation of DNA in the cerebrospinal fluid (CSF). Short interspersed nuclear elements (SINEs) from throughout the genome are PCR-amplified with a single primer pair and the PCR products are evaluated by next generation sequencing. Real-CSF uses machine learning to assess three features from the sequencing data – gains or losses of 39 chromosome arms, focal amplifications, and somatic nucleotide variants. Real-CSF was applied to 282 CSF samples and correctly classified 71 % of 187 cancers and misclassified only 4.2% of 95 non-neoplastic lesions in the brain.

## Introduction

Central nervous system (CNS) neoplasms comprise a heterogenous class of tumors that are either primary, i.e., originate in the brain or spinal cord, or metastatic, i.e., cancers that spread to the CNS from another organ. Approximately 24,500 cases of primary brain cancers occur a year in the United States, with the most common being glioblastoma in adults and medulloblastoma in children ^1^. Metastatic spread to the brain is even more common, accounting for 100,000 cases a year in the United States alone, with lung and breast being the most frequent. Cancers can spread to the brain matter itself, called parenchymal metastases (PM) or to the covering of the brain, known as leptomeningeal disease (LMD).

A pressing clinical challenge is the lack of reliable biomarkers for the diagnosis and monitoring of cancers involving the CNS. The current gold standard is cytology on cerebrospinal fluid (CSF), which has a sensitivity that ranges from 2% to 50%, depending on cancer type ^2^. To achieve maximum sensitivity, cytology requires large (> 10 ml) volumes of CSF, sometimes necessitating serial lumbar punctures ^3^. Magnetic resonance and other imaging procedures cannot reliably distinguish cancer from inflammatory or other non-neoplastic processes and can detect disease only after it has caused anatomic perturbations ^4^. Therefore, biopsy remains the only means for definitively diagnosing CNS neoplasms. Brain biopsies require general anesthesia and hospitalization, are fraught with risks including neurological injury, and carry tremendous financial burden.

There have been several promising types of biomarkers proposed for CNS tumors. Given the relative lack of tumor derived material in the blood (^5 6^), CSF has become an appealing biofluid to explore for diagnosis (^7 8 9 10 11 12^). While CSF sampling is more invasive than venipuncture, it is already part of standard of care for management of several CNS neoplasms, including medulloblastoma, leptomeningeal disease, metastatic spread, and central nervous system lymphomas. In lymphomas, cerebrospinal fluid is routinely sent for cytology and flow cytometry. Though the sensitivity is < 50%, lymphoma patients with CSF positive cytology can proceed directly to chemotherapy and radiation therapy without surgical biopsy.

Liquid biopsies that detect tumor-derived DNA in plasma (circulating tumor DNA, called ctDNA) are now being widely explored to detect and monitor cancers of many types (^13 14 15 16^ and reviewed in ^17^). Implementing analogous tests in CSF is challenging because the quantity of circulating DNA in CSF is considerably lower than in plasma ^5^.

In the current study, we describe our efforts to develop a simple strategy, called Real-CSF, for the diagnosis and monitoring of several of the most common and debilitating brain cancers: glioblastomas, metastatic lesions, lymphomas, and medulloblastomas.

## Results

### Patient Characteristics

Real-CSF uses a single primer pair to PCR-amplify ∼350,000 short interspersed nuclear elements (SINEs) from throughout the genome. As described in Methods, these PCR products of SINEs are assessed by next generation sequencing, and machine learning is used to assess gains or losses of 39 chromosome arms, focal amplifications, and Apparent Somatic Mutations. Note that we did not analyze DNA from normal tissues of the same patients. It is therefore possible that, even after the exclusions detailed in the Methods, a subset of the mutations we detected were rare germline variants rather than true somatic mutations, and we therefore dubbed them “Apparent Somatic Mutations” rather than “Somatic Mutations.”

Two independent cohorts of patients were evaluated in this study: a training set and a validation set. The training set was composed of CSF samples from 92 patients, 37 with GBM, 14 with metastasis from primary tumors outside the brain, 7 with lymphoma, and 34 without cancer. The validation set was composed of CSF samples from 190 patients, 27 with GBM (five of which were pediatric H3K27M diffuse midline gliomas), 52 with metastasis from primary tumors outside the brain, 27 with CNS lymphoma, 23 with medulloblastoma, and 61 without cancer (Fig. 1). Thirteen metastatic samples were previously analyzed using an alternate assay and reported in Naidoo et al. ^18^. The CSF was obtained in almost all cases from lumbar puncture or aspiration from a ventricular catheter placed as part of standard of care. The general demographics of the training and validation set are presented in Supplementary Table S1.

**Figure 1:**
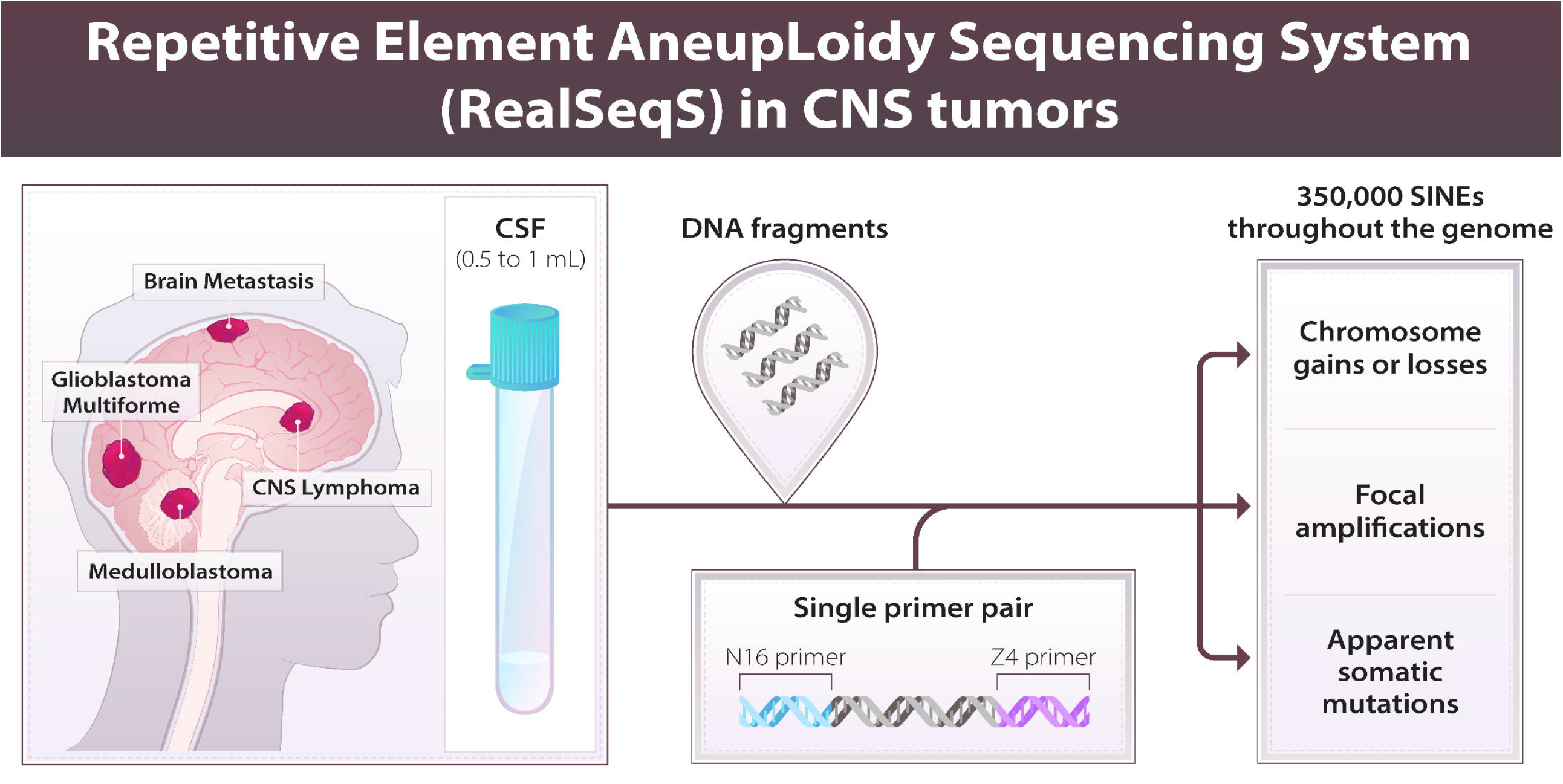
Overview of the Real-CSF approach: Using a single PCR primer to concomitantly amplify ∼350,000 Alu SINE elements spread throughout the genome, Real-CSF uses supervised machine learning to combine large scale chromosome aneuploidies with known focal changes in cancer of the Central Nervous System and mutation burden to detect the presence of cancer.

### Training Set Data

Our goal was to discriminate CSF from patients with and without central nervous tumors. Sensitivity was determined by the fraction of patients with cancer above a given threshold while specificity was determined as the fraction of patients without cancer less than this threshold. We used the training set to determine reasonable thresholds for each of the three parameters - global aneuploidy score, focal amplification Z Scores, and apparent somatic mutations burden – as described in Methods.

For each metric, thresholds were selected based on the largest observed magnitude in the 34 non-cancer training samples. Under this approach, no non-cancer training samples will be positive. Because this is a training set and we are retrospectively selecting an idealize threshold, this does not provide a true estimate of specificity. True sp^19^ecificity should be reported in an independent validation set after predetermining thresholds for positivity. Because the independent validation set contains more samples from more institutions, we anticipate a reduction in specificity. To calculate aneuploidy, Z_w_ scores for each of the 39 non-acrocentric chromosome arms in each sample were calculated (detailed in Methods). These chromosome arm-level Z_w_ scores were then integrated into a single score, called the Global Aneuploidy Score. The Global Aneuploidy Score reflects the likelihood that a sample of interest has gained or lost at least one chromosome, with the magnitude of the score reflecting both the number of chromosomes lost or gained as well as the fraction of cells in which these changes occurred. With n Aneuploidy Score threshold of 0.25, 57% (CI 43% to 70%) of CSF samples from patients with CNS cancers in the training set (51% of patients with GBM, 86% of patients with metastases to the brain, and 29% of patients with lymphomas) scored positive (Fig 2A).

**Figure 2:**
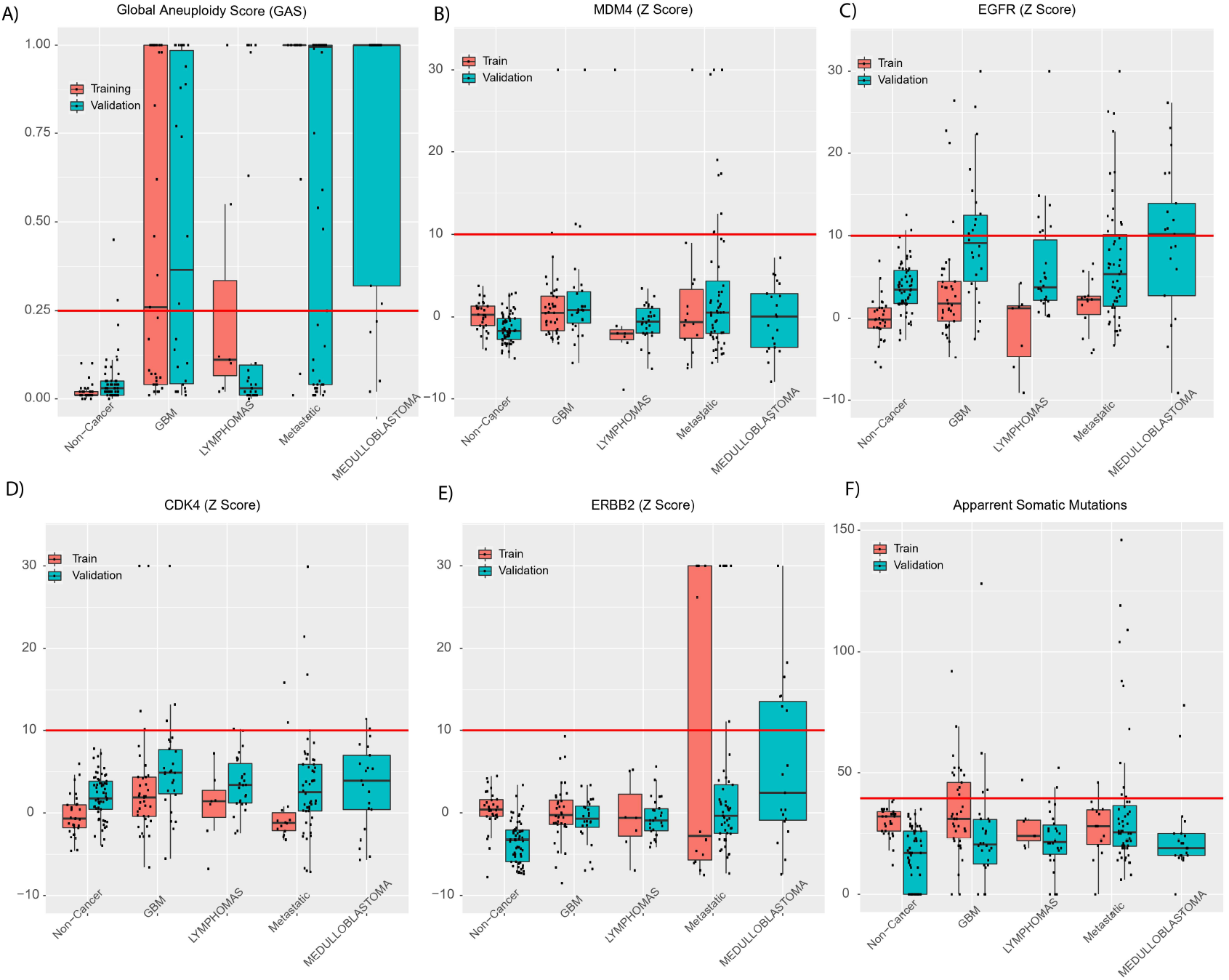
Real-CSF is a multi-analyte test. We illustrate the distribution of all analyte values for each cancer type in both the training and validation cohorts. The threshold for positivity is illustrated by a red horizontal line. A) Global Aneuploidy Score (GAS) B) MDM4 Z Score (Focal Panel) C) EGFR Z Score (Focal Panel) D) CDK4 Z Score (Focal Panel) E) ERBB2 (Z Score) and F) Apparent Somatic Mutations

The Focal Amplification Score is based on a metric called Z_gene_, reflecting the likelihood that a sample harbors an amplification of at least one of four bona fide oncogenes previously shown to be amplified in cancers of the brain and other organs. A Focal Amplification Z Score of 10 was used as the threshold, as explained in Methods. With this threshold, 28% (CI 17% to 41%) of CSF samples from patient with CNS cancers in the training set (24% of patients with GBM, 43% of patients with metastases to the brain, and 14% of patients with CNS lymphomas) scored as positive. Dot plots are graphed in Fig. 2B-E, respectively, and representative examples of data from cases with focal amplifications are shown in Fig. 3. Twenty-eight percent of the CSF samples that did not score positively for aneuploidy had a positive Focal Amplification Score. We selected to the focal thresholds to ensure none of the 34 samples from patients without CNS cancers in the training set were positive for focal amplifications.

**Figure 3:**
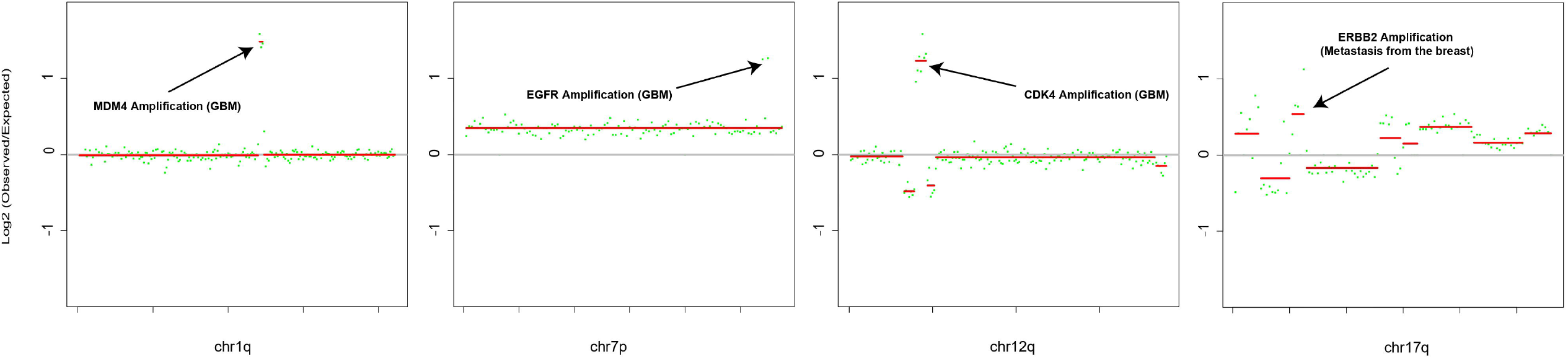
Representative Focal Changes used in Real-CSF. The Real-CSF Focal panel calls focal changes surrounding the following genes: A) 1.5M focal amplification of MDM4 at 1q32.1 (chr1: 203,800,000-205,300,000 hg19) B) 3.5 MB focal amplification of CDK4 at 12q14.1 (chr12: 57,600,000-61,100,000 hg19) C) 1.5 MB focal amplification of EGFR at 7p11.2 (chr7: 54,200,000-55,700,000 hg19) D) 2.5 MB focal amplification of ERBB2 17q12 (chr17: 35,300,000-37,800,000 hg19)

Somatic mutations in a sample have often be used to evaluate whether neoplastic cells contributed to its DNA (as reviewed in ^17 5 20^). Many of these studies use cancer driver genes as the most reliable indicators of neoplasia, because they are known to have functional consequences. However, few if any of the mutations observed in RealSeqS amplicons are likely to have functional consequences because the vast majority of them are outside coding regions of the genome. On the other hand, we previously reported that the number of mutations in repetitive elements is proportional to the number of mutations found by exome sequencing and can be used as a surrogate for the number of somatic mutations, often referred to as “Tumor Mutation Burden” ^21^. We therefore measured the number of Apparent Somatic Mutations (Methods) in RealSeqS data on DNA from CSF. With a threshold of >39, 26% (CI 16% to 39%) of cancers (35% of GBM, 7% of metastatic lesions, 14.3% lymphomas) scored as positive. Dot plots of these scores are graphed in Fig. 2, respectively.

Finally, some of the cancers that did not score positively for aneuploidy did score positively for either focal amplifications or Apparent Somatic Mutations, and vice versa, as noted above and detailed in Table S3. We therefore integrated all three metrics into a single composite score, called Real-CSF. The threshold for Real-CSF was simply a boolean OR gate applied to the thresholds based on its three assay components. In other words, if any of the three assays on a sample were above the threshold defined for positivity for that assay, the sample was scored as positive. Though non-conservative, we believed that this metric would increase sensitivity at the expense of specificity, reasoning that a false positive call would be relatively less deleterious to patients with suspected CNS cancers than a false negative call. In the composite Real-CSF assay 69% (CI 55% to 80%) of cancers (68% GBM, 86% of metastatic lesions, and 43% of lymphomas) scored as positive. Dot plots for Real-CSF data are graphed in Fig. 2.

### Validation Set

The validation set provided an opportunity to independently assess the sensitivity and specificity of RealSeqS in CSF. Note that the validation set included samples from four different institutions, while the samples in the training set were all from only one of these four institutions. This multi-institutional acquisition was designed to minimize confounders previously observed when a classification method based on samples from a single institution is applied to samples from other institutions.

Using the threshold pre-defined by the training set data, 71% of cancers (85% of GBM, 74% of metastatic lesions, 44% of lymphomas, and 78% of medulloblastomas) scored positive in Real-CSF (Fig. 4A). No sample types (GBM, metastatic, lymphomas, non-cancer) present in both the training and validation sets had statistically different detection rates (P>0.05 Two Proportion Z-Test). Interestingly, most samples from patients with medulloblastomas in the Validation set were scored as positive, even though medulloblastoma patients were not represented in the training set (Fig. 2 and Fig. 4). Of the 61 samples from patients without CNS cancers in the validation set, four (6.6%, CI 2.1% to 16.7%) scored positive in Real-CSF.

**Figure 4:**
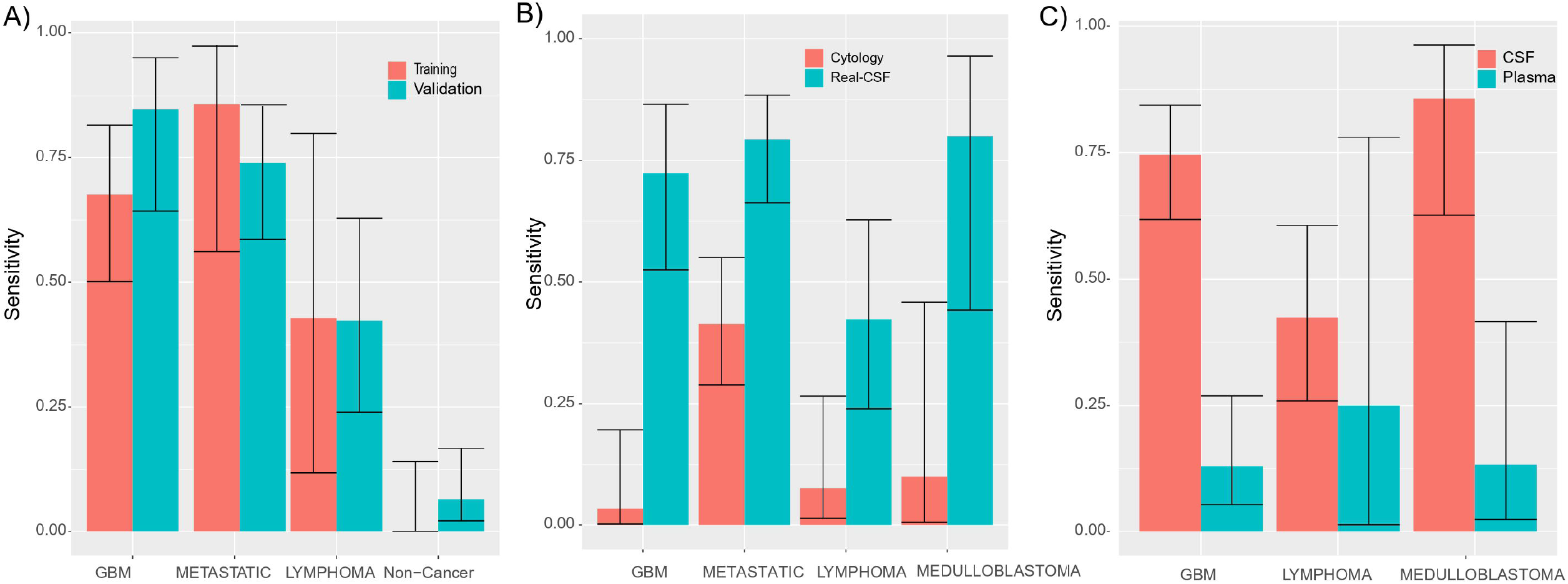
Evaluation of Real-CSF: A) Comparison of performance of Real-CSF in the Training and Validation Partitions. Medulloblastoma is not illustrated because it was not included in the training set, C) Comparison of Performance of Real-CSF to Cytology, Comparison of Performance of Real-CSF in CSF and Plasma.

Of the 92 cancers scoring positively with Real-CSF, 11% were detected by all three of its component assays, 46% by two assays; and 43% by one assay. None of the four false positive samples had more than one assay positive; two were positive for aneuploidy and another two were positive for Focal amplifications (Table S3)

### Comparison to Cytology

CSF cytology is used standard of care for patients with suspected CNS neoplasms. Of the 123 cases in whom cytology was available, 70% were detectable by Real-CSF assay while only 23% were detectable by cytology. In the 28 cancer cases with positive cytology, Real -CSF detected 82% and in the 95 cases with negative or indeterminate cytology, Real-CSF detected 66%. Together, either Real-CSF or cytology were positive in 74% (CI 65% to 81%) of cases (Fig. 4B).

### Survival Analysis

There were sufficient follow-up data to analyze progression free and overall survival in subjects with GBM treated at one of the institutions, JHU. Of the 14 newly diagnosed GBM patients, 10 had detectable levels of CSF-tDNA, while 4 did not. The individuals with detectable levels of CSF-tDNA had an odds ratio of 5.1 (p = 0.02, log rank test, Supplementary Figure 1A) for disease progression when compared to those without CSF-tDNA detection. Of the 29 newly diagnosed and recurrent GBM patients, 20 had detectable levels of CSF-tDNA and 9 had undetectable levels. The cases with detectable CSF-tDNA had an odds ratio of 2.4 for poorer overall survival (p = 0.011, log rank test, Supplementary Figure 1B).

### Analysis of Plasma from Patients with CNS cancers

Given that plasma is much more accessible than CSF, it was of interest to determine how well assays for aneuploidy, focal amplifications, and apparent somatic mutations on plasma DNA compared to those on CSF DNA. We were able to obtain 65 plasma samples from patients with either GBM, lymphomas, or medulloblastomas (Table S4) We also scored 185 non-cancer plasmas to assess specificity and used thresholds identical to those used for the analysis of CSF (Table S4).

Positive Global Aneuploidy Scores were obtained in 14% (CI 6.9% to 25%) of the 65 CNS cancer patients, and none from focal amplifications at a specificity of 98.9% and 99.5% respectively. The apparent somatic mutation count, however, could not reliably distinguish cancers and non-cancers in plasma. The cutoff of > 39 somatic mutations produced a much higher rate of positivity and identified 57.8% of the non-cancers and 67.7% of the CNS cancer plasmas.

Thirty-five of the 65 patients donating plasma samples also had donated CSF (Table S4). In these matched samples, 66% of the CSF samples scored positive in Real-CSF while only 23% of the plasma samples scored positively. There were three (9%) of the 35 cases in which a positive score was observed in plasma but not in CSF, twenty (60%) of the 35 cases in whom a positive score was observed in CSF but not plasma, and five patients in whom positive scores were observed in both plasma and CSF. We conclude that CSF is a more informative source of DNA for assessing aneuploidy and related parameters than plasma in any of the tumor types assessed (P<0.00001, Z Score for 2 Population Proportions, Fig. 4C).

## Discussion

There are many non-neoplastic diseases that present with an imaging abnormality that mimics a neoplastic process. Correctly identifying the lesions as non-malignant is critical, as such patients would not benefit from surgical biopsy and can almost always be managed non-operatively. In contrast, patients with cancer would benefit from immediate consultation with appropriate neurosurgical and oncology specialists.

Biomarkers for distinguishing non-neoplastic from neoplastic lesions, as well as improved imaging techniques, are currently the best hopes for resolving the diagnostic conundrum that exists for thousands of patients with space-occupying lesions of the brain. While a blood-based biomarker would be easier to access, paired analysis of plasma and CSF suggests that CSF is significantly enriched for tumor derived analytes. The ideal biomarker would not require brain tissue, would require only a small amount of CSF, would be simple to interpret, relatively inexpensive, could identify a myriad of cancer types and be done robotically. Real-CSF satisfies all of these criteria and appears to be more informative than cytology, the current gold standard. In addition, Real-CSF shows for the first time the applicability of a multi-analyte approach in CSF, in this case employing copy number changes, sub-chromosomal alterations and apparent somatic mutations for the detection of multiple cancers.

Other groups have demonstrated that low pass whole genome sequencing (WGS) can identify chromosomal copy number alterations in the CSF derived from individuals with select brain cancers (^22 8^;). While WGS has promise, Real-CSF has distinct advantages. It is less expensive, a more simplified workflow, fewer computational requirements and requires minimal starting material.

The ability to identify amplifications and deletions is of increasing importance in neuro-oncology. They can identify potential therapeutic targets and help distinguish different categories of brain cancers.

For example, based on imaging and clinical findings, GBM and lymphoma can have overlapping presentations but face drastically different clinical approaches. Real-CSF has the potential to distinguish between these entities based on patterns of chromosomal alterations and apparent somatic mutations. GBM frequently has a gain on 7p and 7q and losses on 10p and 10q—all infrequently observed in lymphoma. Conversely, lymphoma often has a gain on 18q and very few chromosome arm losses. These chromosomal alterations alone could accurately distinguish 73.0% of the Real-CSF positive GBM and lymphomas in the current cohort. With additional samples, we anticipate the performance will improve and allow accurate identification and classification of other cancer types beyond just those tested in the current study.

There are preliminary data to suggest Real-CSF can be used for the longitudinal monitoring of brain cancers. The first illustrative case is CGLIA 303, a patient with leptomeningeal squamous cell cancer metastasis involving the high cervical spine. The patient was treated with immunotherapy and had a robust response (Supplementary Fig. S2) ^18^. The patient’s initial CSF sample was positive but the second and third samples were negative. Serial MRI scans showed eventual resolution of the enhancement but Real-CSF was able to detect the resolution of disease approximately 8 months prior to radiographic clearance. It is well appreciated that MRI changes lag often months behind biological changes (^23^). CGLIA 301 was a case in which the patient had an MRI that did not show signs of LMD but six weeks later the patient had multiple areas of enhancement suggestive of LMD (Supplementary Fig. S3). In both cases, Real-CSF was able to accurately predict disease status before radiographic findings emerged.

The standard of care treatment for glioblastoma includes concomitant chemo and radiation therapy. Discerning true disease recurrence from treatment related changes (pseudoprogression) on imaging can be very challenging ^19^. Frequently, individuals are taken to surgery for pathological confirmation of disease status and approximately 30% of cases will have only treatment effect on histological examination ^24^. If a biomarker was able to discern active disease from pseudoprogression, it may obviate the need for surgery in select patients. GLIA 566 is a case that suggests Real-CSF may be able to serve as such a biomarker. The subject had completed treatment with temozolomide and radiation therapy but was found to have progressive enhancement on MRI and was taken to surgery to distinguish tumor progression from treatment effect (Supplementary Figure S4). Real-CSF accurately demonstrated active disease, which was confirmed on pathological examination. In the future, testing with an assay such as Real-CSF may help accurately identify disease status without the need for neurosurgical intervention.

For a CSF biomarker to be clinically viable, it must be able to distinguish neoplastic from non-neoplastic conditions. For the first time, we test the performance of an assay designed to detect tumor derived DNA in a sizeable cohort (95) of CSF samples from individuals without cancer but with a variety of other neurological conditions. These control samples represent inflammatory, autoimmune, degenerative, congenital and vascular conditions affecting the brain and central nervous system. The specificity in the Validation set was 93%, which is comparable to CSF cytology, the current gold standard (^2^).

While the sample size was small and needs to be confirmed on larger datasets, CSF-tDNA detection may correlate with progression free and overall survival in GBM, a finding that others have reported in the literature ^25 26^;. This suggests that cancers that shed tumor derived material into the CSF could represent a more aggressive phenotype, information that may be used to help guide clinical decision making.

Though Real-CSF is a step in the right direction, it is not perfect. While specificity could be enhanced, perhaps more importantly, its sensitivity could be improved; in the Validation set, it was able to detect only 85% of GBM, 74% of metastatic lesions, 44% of lymphomas, and 78% of medulloblastomas. One opportunity to augment sensitivity in future studies would be to analyze larger volumes of CSF. The current study utilized 1 ml or less of CSF, which for comparison is only ∼10% of what is utilized currently for cytological examination. While these experiments are the first to demonstrate utility with a multi-analyte approach in CSF, they are all genetic in origin. Integrating additional analytes such as proteins, microRNA or epigenetic markers, all of which have shown promise in brain cancers ^27 28 29 30^, and reviewed in ^12^ may further improve performance.

Our study has several other limitations. Although the total number of samples in the manuscript is large when compared to previously published CSF biomarker studies, confidence limits for subtypes of cancers could be narrowed by studying more patients. Second, the optimal way to define thresholds for Real-CSF would be to use a second set rather than from the Training set used to develop the initial models. These thresholds could then be used to establish sensitivity and specificity in a third, independent cohort. We did not have sufficient number of samples to do this in the current study,.

A third limitation is that this study was retrospective in nature. A large, prospective study will be required to progress Real-CSF as a diagnostic tool. In such a future study, the performance of both cytology and Real-CSF could be assessed in all samples, and we expect that both could be used together to increase sensitivity while retaining specificity for brain cancers. The results reported here establish the conceptual and practical foundation for such a future study, which could have a substantial impact on standard of care.

## Methods

### Sample acquisition

This study was cross-sectional in design. Patients were recruited as part of an Institutional Review Board-approved, multi-institutional study to develop biomarkers for central nervous system tumors using cerebrospinal fluid. The four institutions involved (Johns Hopkins, University of Michigan, Penn State University, Children Tissue Brain Tumor Tissue Consortium (CBTTC)) are tertiary centers that care for patients referred for management of central nervous system tumors. In general, patients underwent sampling on the day of enrollment and only tumors with radiographic confirmation with contrast enhanced MRI were included in the study. Radiographic findings of disease were based on the findings of a board certified-neuroradiologist at each site. In total there were 282 samples collected for this study. Pathologic diagnosis for all cases was verified by board-certified neuropathologists at the site of enrollment. Patients whose diagnosis was GBM, CNS lymphoma, or metastasis from outside the brain comprised the true positive subset. Patients who were not diagnosed with any neoplastic disease comprised the true negative set. We were also able to assess plasma samples from 65 cancer patients of which 35 had matched CSF for comparison purposes.

Samples were pre-specified into training and validation cohorts based on the sample source and the time in which the sample was available for evaluation at Johns Hopkins. An initial batch of samples from Johns Hopkins were labeled as training samples. To reduce potential cohort biases and overfitting from machine learning, all samples from the Penn State University, CBTTC, and the University of Michigan were labeled as validation samples. The remaining Johns Hopkins samples not evaluated in the initial batch of samples were included in the validation set.

### DNA purification

CSF was frozen in its entirety at -80 ^⍰^C until DNA purification, and the entire volume of CSF (cells plus fluid) was used for DNA purification. The amount of CSF used for purification ranged from 0.5 to 1 mL. CSF and plasma DNA (from 1 mL of plasma) was purified from healthy individuals and patients using Biochain reagents according to the manufacturer’s instructions (catalog #K5011625MA).

### Real-CSF

A single primer pair was used to amplify ∼350,000 short interspersed nuclear elements (SINEs) spread throughout the genome ^31^.PCR was performed in 25 uL reactions containing 7.25 uL of water, 0.125 uL of each primer, 12.5 uL of NEBNext Ultra II Q5 Master Mix (New England Biolabs cat # M0544S), and 5 uL of DNA. The cycling conditions were: one cycle of 98°C for 120 s, then 15 cycles of 98°C for 10 s, 57°C for 120 s, and 72°C for 120 s. Each sample was assessed in eight independent reactions, and the amount of DNA per reaction varied from ∼0.1 ng to 0.25 ng. A second round of PCR was then performed to add dual indexes (barcodes) to each PCR product prior to sequencing. The second round of PCR was performed in 25 uL reactions containing 7.25 uL of water, 0.125 uL of each primer, 12.5 uL of NEBNext Ultra II Q5 Master Mix (New England Biolabs cat # M0544S), and 5 uL of DNA containing 5% of the PCR product from the first round. The cycling conditions were: one cycle of 98°C for 120 s, then 15 cycles of 98°C for 10 s, 65°C for 15 s, and 72°C for 120 s. Amplification products from the second round were purified with AMPure XP beads (Beckman cat # a63880), as per the manufacturer’s instructions, prior to sequencing. Sequencing was performed on an Illumina HiSeq 4000. The sequencing reads from the 8 replicates of each sample were summed for bioinformatic analysis. The average number of the summed, uniquely aligned reads was 10.5 million (interquartile range, 8.0-12.7 million). Samples with < 2.5M reads were excluded on the basis of previously described metrics ^31^. The bioinformatic methods and pipeline used to process the raw sequencing data are available at (https://zenodo.org/record/3656943#.YaZZCdDMKUk).

### Chromosome Copy Number Alterations CSF

The copy number alterations for CSF samples were calculated using in the following protocol:

1. Generate a euploid reference panel using 15 non-cancer CSF samples from the pre-defined training set.
2. Aggregate and sum the read depth into 5,344 non-overlapping autosomal 500-kb intervals.
3. Perform PCA normalization on the 500-kb intervals. PCA normalization mitigates the impacts of highly correlated outlier chromosome regions that arise from NGS artifacts like GC bias. A full detailed step by step mathematical description and pseudocode can be found in the Supplementary Text of Douville et al. 2020 ^31^.
4. Segment the chromosome arm using the circular binary segmentation algorithm (CBS) ^32^
5. Aggregate the 500-kb intervals across the chromosome arm and calculate the statistical significance across the length of the chromosome arm (Z_w_).
6. Repeat this protocol for all chromosome arms.
7. Train a supervised machine learning algorithm that generates a Global Aneuploidy Score (GAS) to discriminate between aneuploid and euploid samples. The predictive features of the model are the 39 chromosome arms (Z_w_). The training examples were 3,999previously published plasma samples. The negative class of 1348 presumably euploid samples were taken from individuals without cancer. The positive class was taken from 2651 aneuploid samples across 8 different cancer types. We specifically built a support vector machine (SVM) and trained the model with the e1071 package in R, using a radial basis kernel and default parameters.
8. Score the test sample using the supervised-machine learning model from Step 7.
9. The full bioinformatic pipeline is available at: https://zenodo.org/record/3656943#.YaZZCdDMKUk.

### Chromosome Copy Number Alterations Plasma

In order to identify copy number alterations in plasma we repeated the steps from above but made one key change. We reconstructed the euploid reference panel using a set of 1,500 euploid plasma samples. We then repeated the step-by-step protocol as above to calculate the statistical significances for each arm and generate Global Aneuploidy Scores.

### Focal Amplifications

We chose a set of genes for evaluation of focal amplifications based on data reported in CNS cancers from The Cancer Genome Atlas (TCGA) ^33^. The four genes chosen were MDM4, EGFR, CDK4, and HER2 (genomic coordinates in Table S2). RealSeqS amplicons overlapping these five genes plus an additional 1 MB flanking each side of the gene were identified. The summed read counts (Observed_gene_) across these amplicons was then determined for each sample.

The protocol to calculate the Z score for each gene was calculated as such:

For the euploid reference panel:

1. For all samples in the reference panel, normalize each locus by dividing by the total autosomal sequencing depth. This enables samples with varying amounts of coverage to be directly comparable.
2. Aggregate the read depth across the gene of interest for each sample.
3. Estimate the average read depth across the euploid reference panel (µ_gene_). For each test sample:
4. Calculate the total autosomal sequencing depth (Coverage)
5. Multiply (µ_gene_) by the observed coverage to estimate the expected number of reads across the gene of interest (λ_gene_) given the coverage. We assume count data follow a Poisson distribution.
6. Aggregate the read depth across the gene of interest (Observed_gene_)
7. Calculate the statistical significance

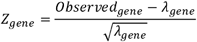

This protocol was followed for both CSF and plasma samples. The only difference between CSF and plasma was the euploid reference panel used to generate the expected depth for each gene. We listed these estimates for both CSF and plasma in Table S2.

### Apparent Somatic Mutations

Adapters were trimmed using cutadapt2 and aligned to the genome using Bowtie2 ^34^. Potential somatic mutations were identified using Mutect2 default parameters ^35^. Multi-allelic, poor mapping quality, poor base-quality, and clustered variants were discarded. Only autosomal chromosomes were considered and an allele frequency cutoff of <0.35 was used to exclude germline variants because we assumed that neoplastic cells contributed <20% of the DNA to the entire DNA purified from CSF samples. Samples in which the Q30 <75 (all cycles) were excluded for mutation analysis because of the increased likelihood of base-calling errors. All samples were quantified with qPCR and samples with <0.03 ng/uL of purified DNA were not used for mutation analysis, also because of the increased likelihood of artifactual mutations in such samples ^36^. We then counted the total number of single base substitution mutations observed in RealSeqS data for each sample.

## Data Availability

The bioinformatic methods and pipeline used to process the raw sequencing data are available at (https://zenodo.org/record/3656943#.YaZZCdDMKUk).

---

**Figure S1:** Progression free (A) and overall survival (B) in subjects with GBM treated at one of the institutions.

**Figure S2:** A) MRI scan CGLIA 303 B) MRI scan CGLIA 303 (6.x4 mm) C) MRI scan CGLIA 303 (6.x 2 mm) D) MRI scan CGLIA 303 (negative)

**Figure S3:** A) CGLIA 301 – No evidence of LMD from breast cancer B) CGLIA 301– Positive LMD from breast cancer

**Figure S4:** A) GLIA 566 post-contrast MRI scan demonstrating enhancement that increased over time B) raising the question of tumor recurrence versus treatment effect.

## Acknowledgements

We would like to thank our patients for their bravery and willingness to participate in research studies. This work was supported by The Virginia and D.K. Ludwig Fund for Cancer Research, The Sol Goldman Sequencing Facility at Johns Hopkins, Burroughs Wellcome Career Award for Medical Scientists, Thomas M Hohman Memorial Cancer Research Fund and Alex’s Lemonade Stand Foundation. In addition, the work was supported by NIH grants RA37 CA230400, U01CA230691 and R21NS113016. This research was conducted using data or samples made available by The Children’s Brain Tumor Network (formerly the Children’s Brain Tumor Tissue Consortium).

