## Supplementary figures and images for "Seq-ing the SINEs of Central Nervous System Tumors in Cerebrospinal Fluid DNA"

### Figure S1

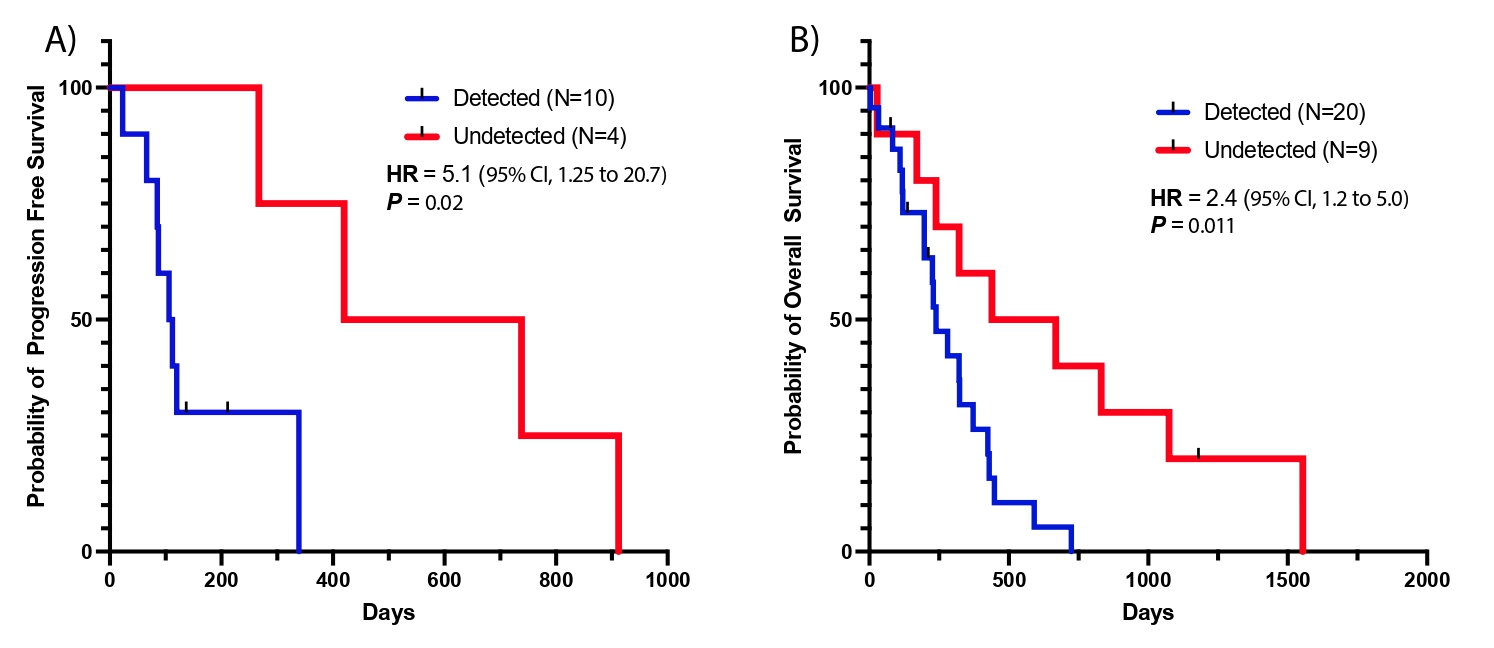

### Figures S2-4

#### Slide 1
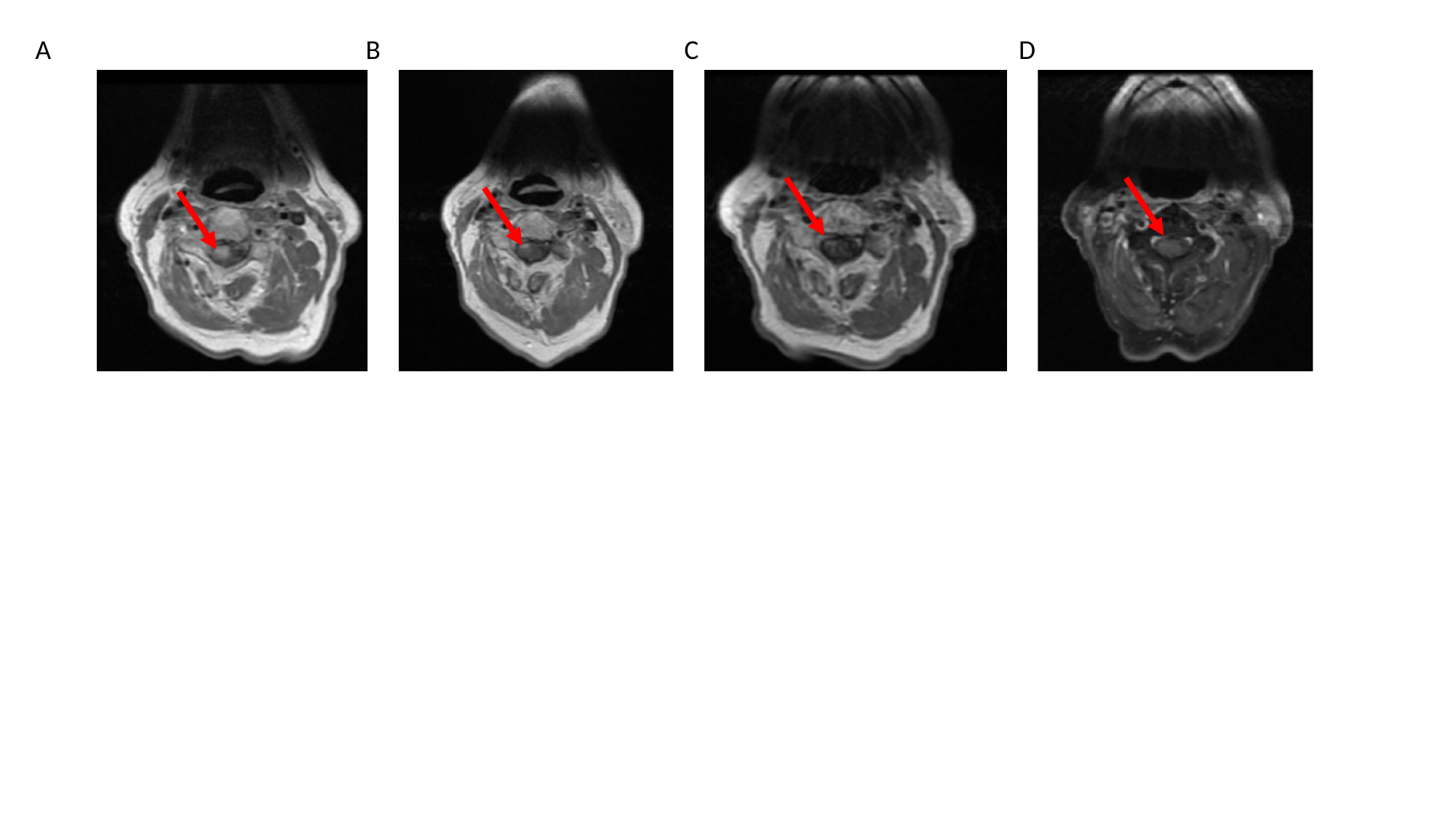

A
B
C
D

#### Slide 2
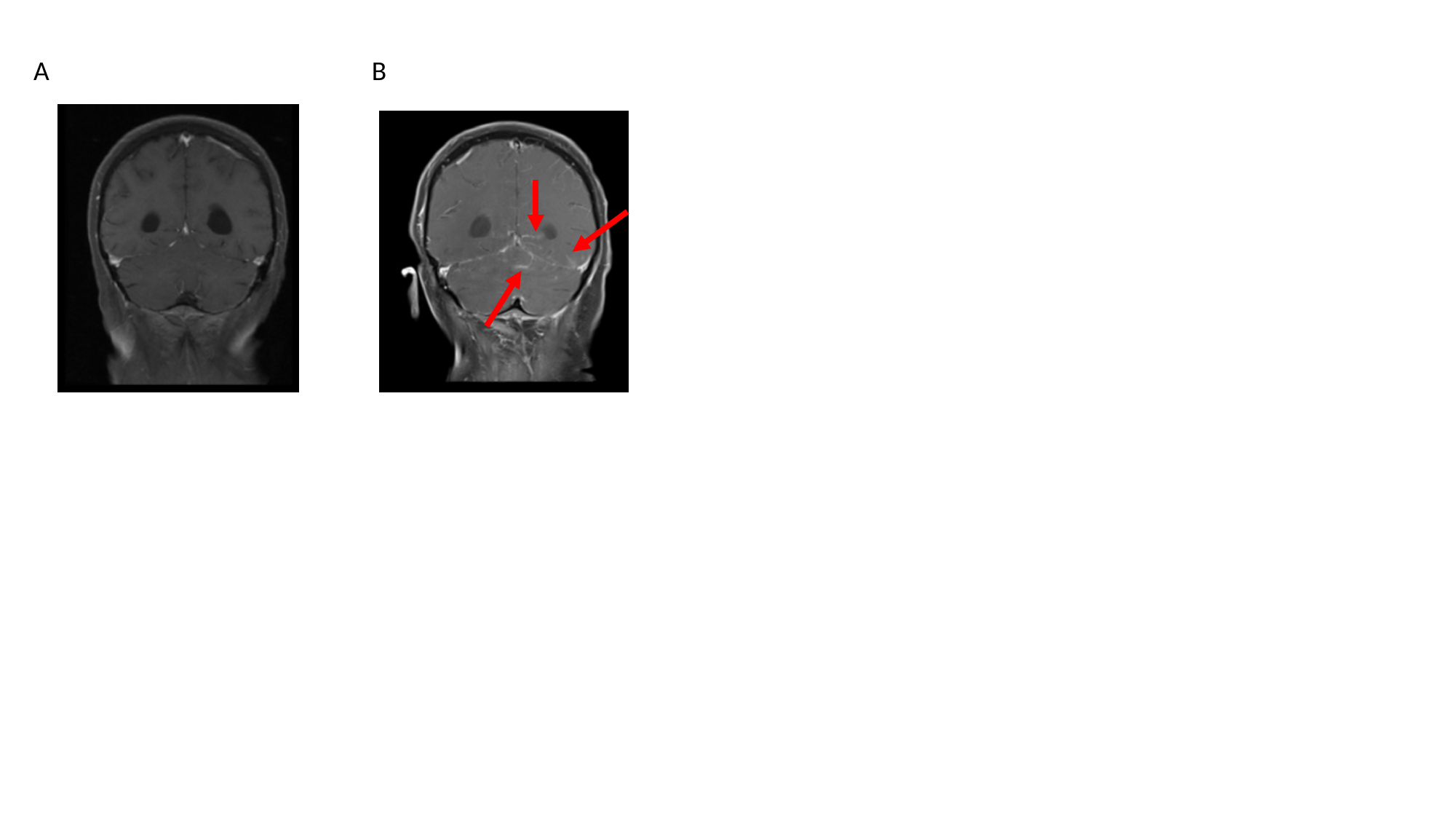

A
B

#### Slide 3
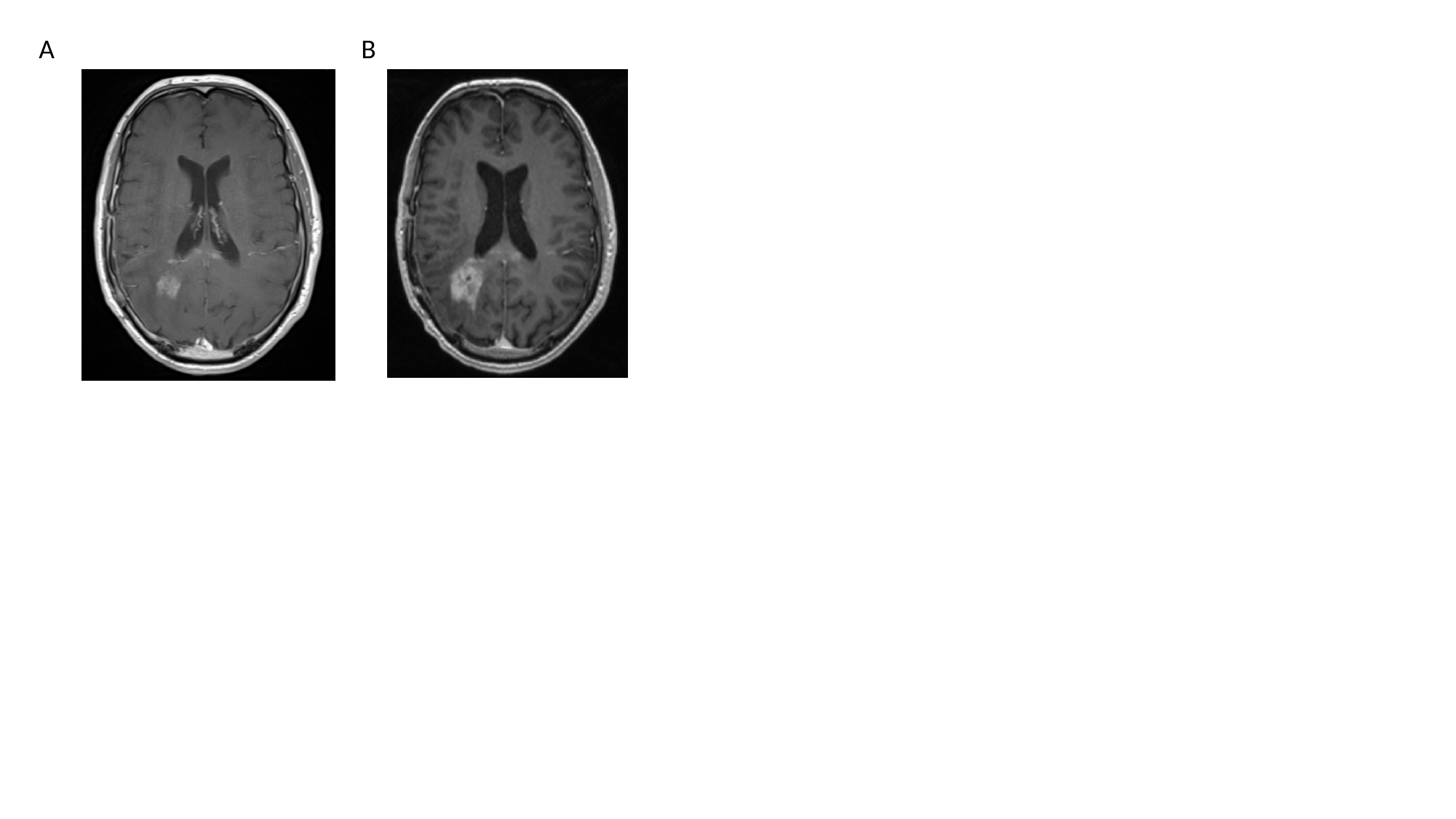

A
B
